# Mortality from COVID-19 in 12 countries and 6 states of the United States

**DOI:** 10.1101/2020.04.17.20069161

**Authors:** Patrick Brown, Kamal Rai, Carlo La Vecchia, Peter S Rodriguez, Kathleen Qu, Mathew G Brown, Hwashin Hyun Shin, Xuyang Tang, Leslie Newcombe, Wilson Suraweera, Craig Schultz, Isaac Bogoch, Hellen Gelband, Nico Nagelkerke, Prabhat Jha

## Abstract

**Importance:** Reliable estimates of COVID-19 mortality are crucial to aid control strategies and to assess the effectiveness of interventions.

**Objective:** Project COVID-19 mortality trends to October 1, 2020, in 12 countries or regions that constitute >90% of the global COVID-19 deaths reported as of April 12, 2020.

**Design, Setting, and Participants:** The Global COVID-19 Assessment of Mortality (GCAM) is an open, transparent, and continuously updated (www.cghr.org/covid) statistical model that combines actual COVID-19 mortality counts with Bayesian inference to forecast COVID-19 deaths, the date of peak deaths, and the duration of excess mortality. The analyses covered a total of 700 million population above age 20 in 12 countries or regions: USA; Italy; Spain; France; UK; Iran; Belgium; a province of China (Hubei, which accounted for 90% of reported Chinese deaths); Germany; the Netherlands; Switzerland; and Canada; and six US states: New York, New Jersey, Michigan, Louisiana, California, and Washington.

**Results:** Forecasted deaths across the 12 current high-burden countries sum 167,000 to 593,000 (median 253,000). The trajectory of US deaths (49,000-249,000 deaths; median 86,000)—over half of which are expected in states beyond the initial six states analysed in this study—will have the greatest impact on the eventual total. Mortality ranges are 25,000-109,000 (median 46,000) in the UK; 23,000-31,000 (median 26,000) in Italy; 21,000-37,000 (median 26,000) in France and 21,000-32,000 (median 25,000) in Spain. Estimates are most precise for Hubei, China—where the epidemic curve is complete—and least precise in California, where it is ongoing. New York has the highest cumulative median mortality rate per million (1135), about 12-fold that of Germany. Mortality trajectories are notably flatter in Germany, California, and Washington State, each of which took physical distancing and testing strategies seriously. Using past country-specific mortality as a guide, GCAM predicts surge capacity needs, reaching more than twice existing capacity in a number of places., In every setting, the results might be sensitive to undercounts of COVID-19 deaths, which are already apparent.

**Conclusion and Relevance:** Mortality from COVID-19 will be substantial across many settings, even in the best case scenario. GCAM will provide continually updated and increasingly precise estimates as the pandemic progresses.

The coronavirus disease (COVID-19) pandemic has already caused over 115,000 deaths, with global deaths doubling every week.^1-3^ Mortality is less biased than case reporting, which is affected by testing policies. However, the daily reporting of COVID-19 deaths is already known to undercount actual deaths, varying over time and place.^4-6^

Reliable estimates of total COVID-19 mortality, the date of peak deaths, and of the duration of excess mortality are crucial to aid responses to the current and potential future pandemics. We have developed the Global COVID-19 Assessment of Mortality (GCAM), a statistical model to project COVID-19 mortality trends to October 1 2020 in 12 countries or regions that constitute >90% of the global COVID-19 deaths reported as of April 12^th^. We report also on six US states that account for 70% of the American totals to date (Supplementary Appendix).^1^ We quantify the COVID-19 mortality trajectory ranges in each setting. A semi-automated website (www.cghr.org/covid) provides daily updates. GCAM is open, transparent, and uses a reasonably simple method that employs publicly reported mortality data to make plausible projections. The method is designed to improve as more mortality data become available over longer time periods.

## METHODS

The goal of the University of Toronto’s GCAM is to provide medium-term mortality forecasts. For each country, US state, and for Hubei Province in China, GCAM uses non-linear regressions to fit a five parameter model to the observed daily mortality counts. We used Bayesian inference, via the algorithm from Carpenter et al.,^7^ of several thousand posterior (“prior”) probabilities of potential epidemic curves for each country. The model builds on observed daily mortality counts for places that have already reached the peak of their epidemic (such as Hubei, China), or are near their peak (e.g. Italy and Spain), where the posterior probabilities are less relevant. For the majority of settings where the epidemic is still accelerating, death counts are currently less predictive, since a variety of curves can fit any particular series of increasing counts. In these cases, GCAM relies on Bayesian prior distributions, which are quantifications of expert knowledge of the underlying process expressed as probability distributions for model parameters. Bayesian prior distributions allow use of past epidemiological knowledge from China where COVID-19 has already declined, as well as the mortality trajectories of diseases caused by similar coronaviruses causing Severe Acute Respiratory Syndrome (SARS)^8^ or the Middle East Respiratory Syndrome (MERS).^9^

## Mortality Data Sources

We systematically collected COVID-19 mortality data for all countries with more than 100 deaths. We examine the top ten countries ranked by the total number of COVID-19 deaths as of April 12^th^: USA; Italy; Spain; France; UK; Iran; Belgium, Hubei province, China (as this accounted for 90% of reported Chinese deaths); Germany and the Netherlands. We also included Switzerland, Canada and the US States of New York, New Jersey, Michigan, Louisiana, California, and Washington. We collated daily mortality data (where reported by age and sex) from the World Health Organization (WHO), country reports,^10^ and from online data collections.^1-3^ Definitions of COVID-19 mortality vary across countries. The WHO has created a new International Classification of Diseases, 10^th^ edition, ICD-10 code, U07.2,^11^ but this has been applied inconsistently across countries. In some countries (e.g., the Netherlands), only deaths of people testing positive for SARS-CoV-2, the causative agent for COVID-19, are counted as definitely due to the virus.^12^ We deemed each setting’s epidemic to have begun on the day after the first death is recorded. We forecast deaths until October 1, 2020, but most deaths will have occurred by July 1.

To enable contrasts with current COVID-19 mortality (which is nearly always from acute respiratory failure), we examined the WHO Mortality database^13^ to quantify the average baseline pneumonia and influenza deaths during 2015-17. For the US, we used the Centers for Disease Control and Prevention (CDC) mortality database,^14^ and for China, we used the Global Burden of Disease.^15^ We obtained age- and sex-specific population denominators from the United Nations Population Division,^16^ and for Hubei, China, and for US states, from national vital statistics.^14,17^

### Statistical modelling of mortality

We assumed the mean number of daily deaths for each setting to follow the shape of a skew-normal density function, which resembles a typical “bell-shaped” normal distribution with added skewness to permit an epidemic to rise fast but fall slowly (as has already been observed with COVID-19 deaths).^1-3^ The observed deaths *Y*_*i*_(*t*) in region *i* on day *t* follows a negative binomial distribution with mean *λ*_*i*_(*t*) and overdispersion parameter *τ*, or more specifically:

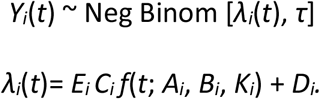

The *E*_*i*_ are age and sex standardized death counts, obtained from each country’s population data and age-specific mortality risk calculated from Italian data on 29 March 2020.^18^ The model parameters which determine the shape of the epidemic curve are: *C*_*i*_, the expected total deaths relative to the Italian rates on 29 March; *A*_*i*_, the location of the peak of the epidemic; *B*_*i*_, the duration of the epidemic (roughly the standard deviation); *K*_*i*_, the skewness parameter specifying the steepness of the growth phase of the epidemic relative to the decline phase; and *D*_*i*_, a ‘spark’ term allowing for occasional deaths outside the range of the main epidemic.

We used the Markov-Chain Monte Carlo algorithm within the Stan software.^7^ From each posterior sample of the model parameters, we computed the intensity *λ*_*i*_(*t*) and simulated potential future death counts from the negative binomial distribution. We calculated predictions envelopes, which contain the entire true epidemic curve from the samples by applying the methodology of Myllymäki et al.^19^ We chose probability distributions to generate simulated epidemic curves consistent with data from Italy and Hubei, China,^1-3^ and with SARS and MERS outbreaks.^8,9^ The Supplementary Appendix provides the prior distributions for all parameters, as well as simulated epidemics for a population with Italy’s demographic structure. As model validation, we fitted the model to training data of deaths on or before March 20 and compared the projections with data observed up to April 12.

For each country, we present the actual mortality daily totals, projected daily and totals, the date of peak mortality and derive credible intervals (CI), at various probability levels. We calculated age-specific mortality rates at age 20 years or more for influenza and pneumonia, and present totals for both sexes, as COVID-19 deaths are only rarely reported in children and adolescents.^20^

## RESULTS

Figure 1 shows the results for six European countries with over 2,000 COVID-19 deaths to date and the USA. Each blue dot indicates daily actual deaths. The dark line is the median model projection and the red shaded area represents credible intervals. Mortality has already peaked in Italy and Spain (March 29 and April 1, respectively). The Netherlands and France next peaked on April 5 and 7, respectively, followed by Belgium (April 9) and Germany (April 13). The USA and UK will reach their peaks on April 18 and 21, respectively. Figure 2 provides these results for Hubei, China and Iran, where peaks occurred earlier, on February 12 and March 28, respectively. The peak date for Switzerland is April 7, and, with less certainty, for Canada, May 4. For the US states, Louisiana has likely peaked first (April 8), followed by Washington state (April 10), New York and Michigan (April 11), New Jersey (April 14), and with less certainty, California (April 14). By July 1, all epidemics will be effectively over.

**Figure 1:**
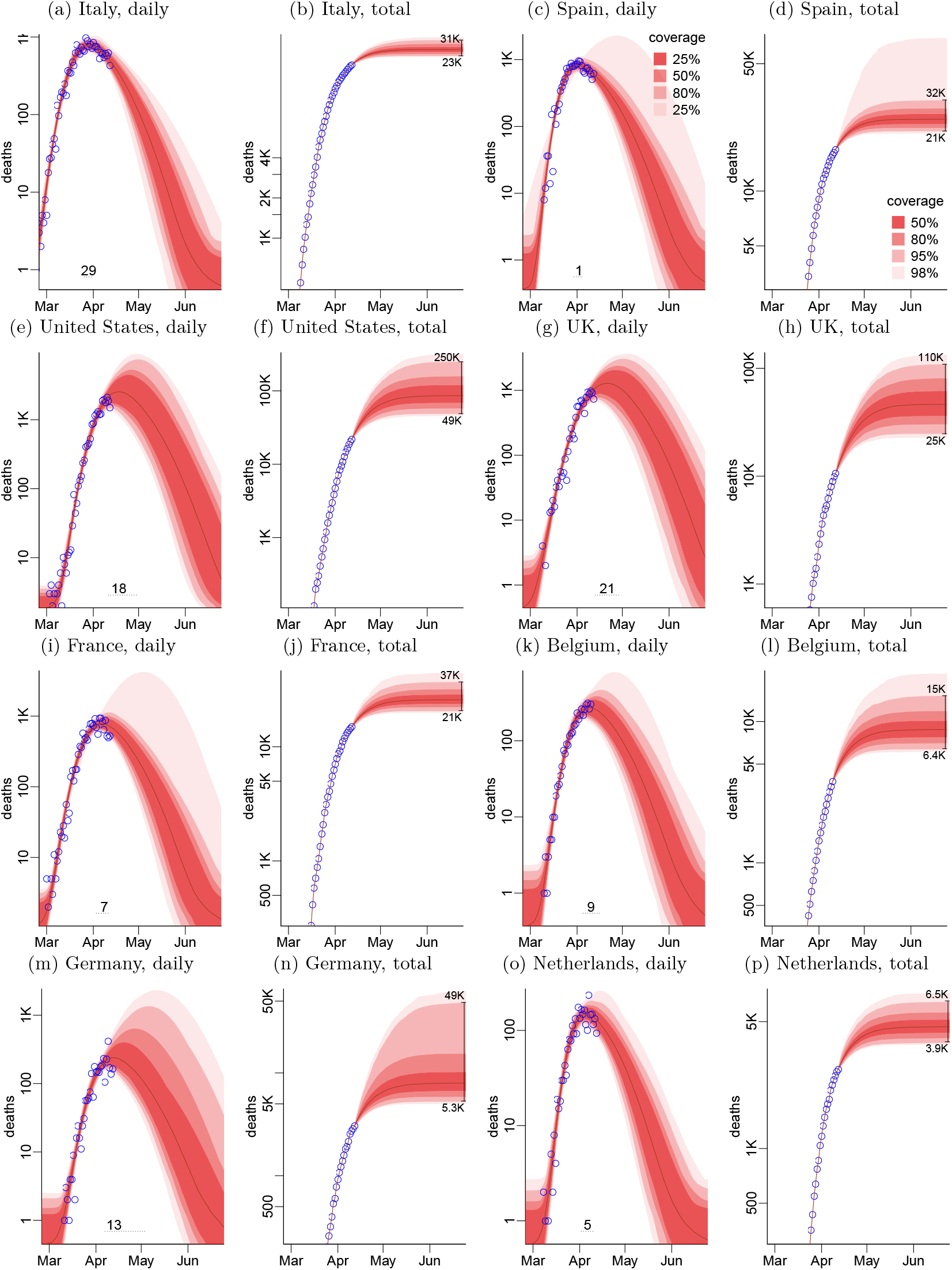
Data and projections for daily and cumulative deaths in the most affected countries. Prediction envelopes (daily are global, cumulative are pointwise) are shaded regions, and observed data are blue plotting symbols.

**Figure 2:**
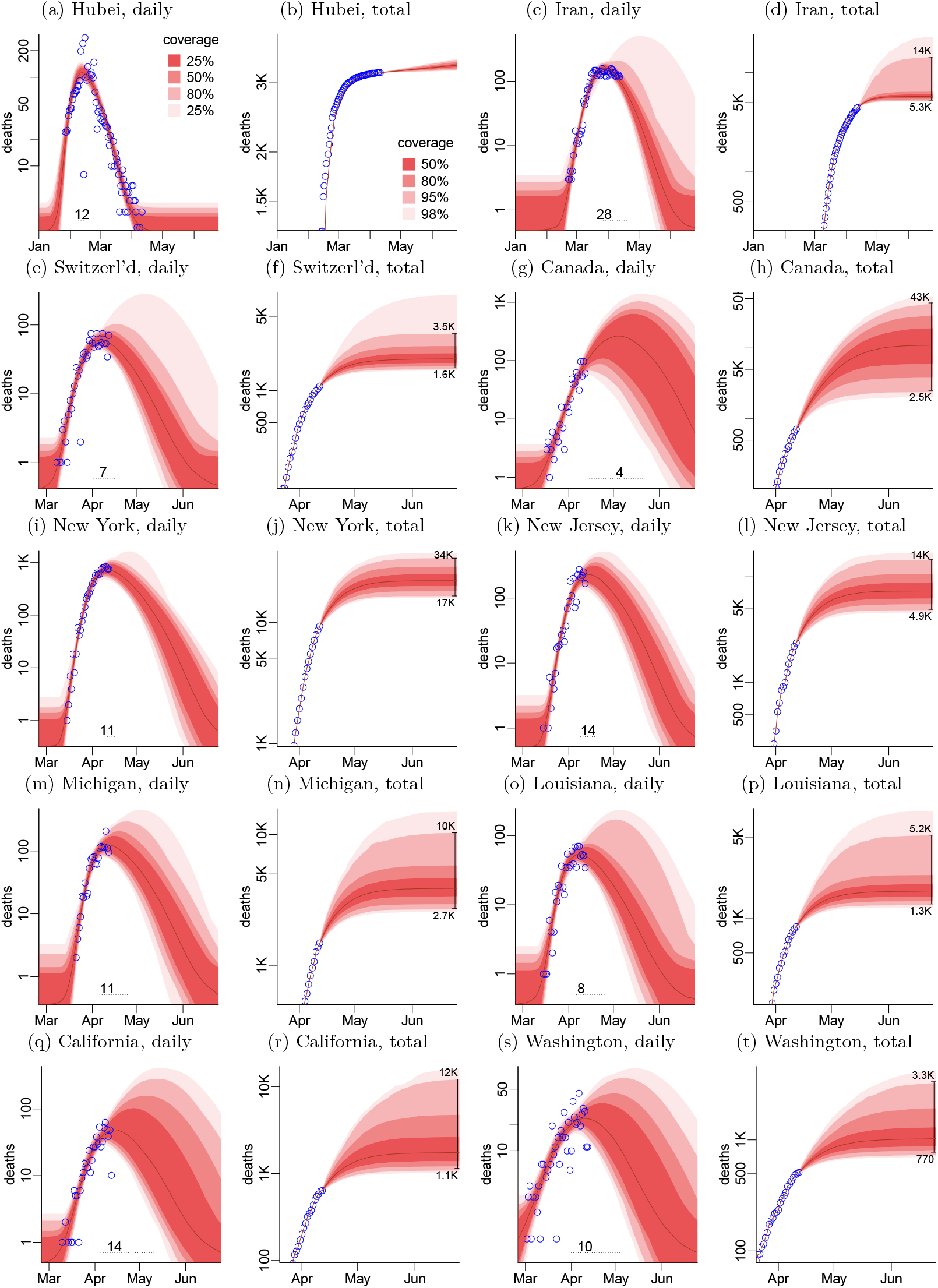
Data and projections for daily and cumulative deaths in various countries and subnational units. Prediction envelopes (daily are global, cumulative are pointwise) are shaded regions, and observed data are blue plotting symbols.

Current COVID-19 deaths, the credible ranges of forecasted deaths, the death rate per 1 million (M) population, and the percentage of national deaths over a whole year for all 18 countries or states/provinces are provided in Table 1. The USA can expect 49,000-249,000 deaths (median 86,000). Mortality ranges are 25,000-109,000 (median 46,000) in the UK; 23,000-31,000 (median 26,000) in Italy; 21,000-37,000 (median 26,000) in France and 21,000-32,000 (median 25,000) in Spain. Ranges for other countries are as shown. Collectively, across the 12 countries, the range of deaths is 167,000 to 593,000 (median 253,000). The lowest value would be approximately a 50% increase in COVID-19 deaths over totals as of April 14 in these countries. The maximum would represent more than 500% increase of deaths. As a proportion of all annual mortality, the median estimate of excess COVID-19 mortality ranges from 1.0% in Hubei to 8.1% in Belgium. Median deaths in the USA and the UK represent 3.2% and 7.8%, respectively of total non-COVID-19 deaths in 2020.

**Table 1:**
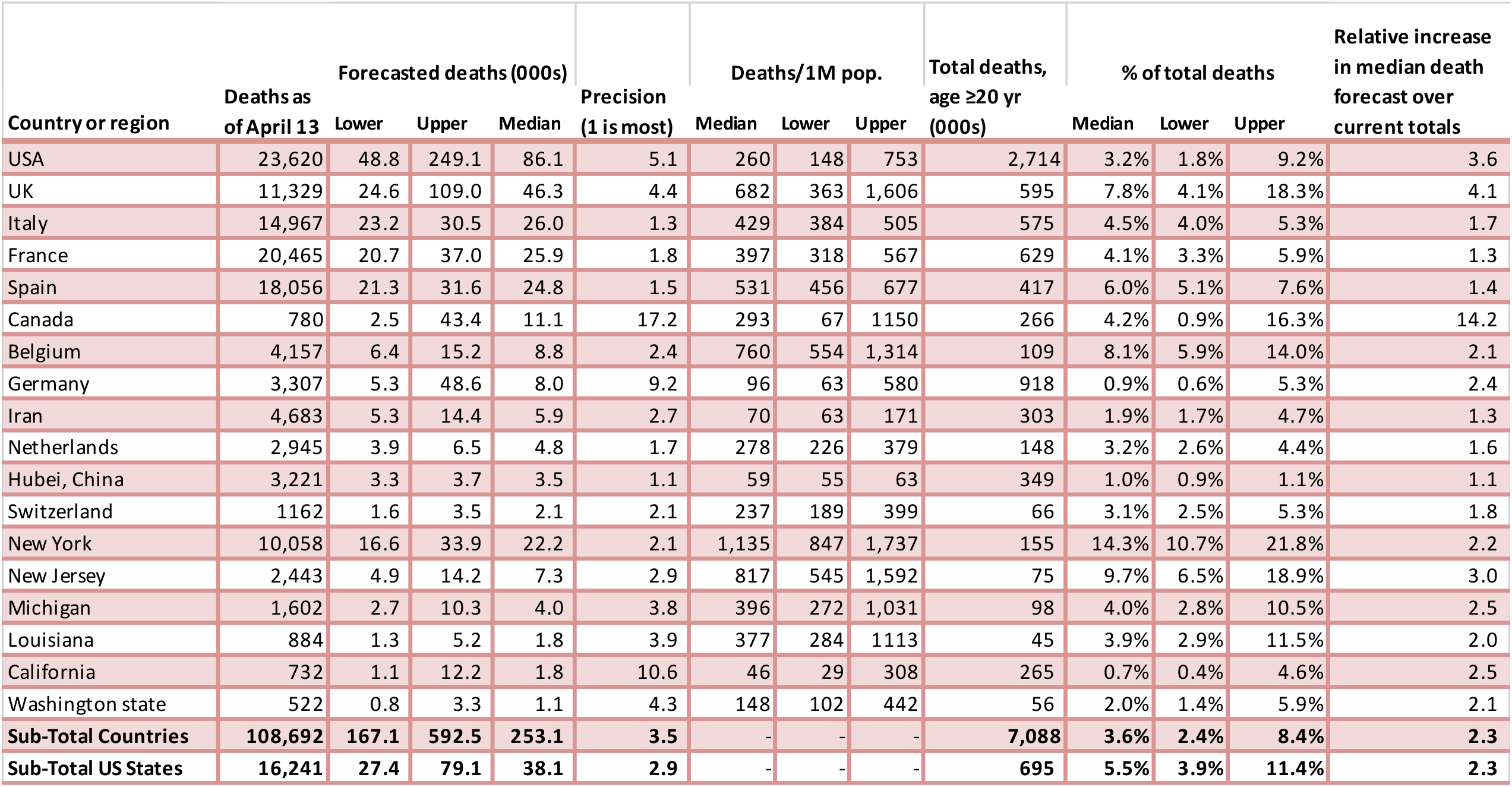
Current COVID-19 deaths, forecasted death ranges, precision estimate, deaths per million and proportion of COVID-19 to total deaths in 12 high-burden countries and six US states.

The column called precision represents the ratio of upper to lower uncertainty estimates. The most precise estimates are in settings where the epidemic has peaked or nearly peaked: Hubei, China (1.1), Italy (1.3), and Spain (1.5). The least precise estimates are for Canada (11.1) and California (10.6). The USA and the UK show moderate precision 5.1 and 4.4, respectively.

Table 2 provides an estimate of the maximal daily totals, ranging from median values of 4,787 in the USA to 114 in Switzerland, and within the USA, individual states ranging from 1,303 in New York to 49 in Washington state. We used the average annual deaths in previous years from pneumonia and influenza in both sexes above age 20 years as a crude indicator of existing hospital and ventilator capacity. We compare these totals against the median COVID-19 deaths, and this ratio provides an estimate of the “surge” capacity needed to deal with the excess COVID-19 deaths. The highest ratios are in France and Spain (each 2.1). We estimate the number of days in which COVID-19 deaths will exceed the daily death total of pneumonia in earlier years to be highest in Canada at 73 (39-104) with other settings averaging 62 days (48-83).

**Table 2:** Peak dates (and credible intervals), maximal deaths per day, comparison with current annual pneumonia deaths and days of excess COVID-19 mortality above pneumonia deaths in 12 high-burden countries and six US states.

| Country or region | Peak date |  |  | Max deaths/day |  |  | Average annual pneumonia deaths in recent years age ≥20, (000s) | Ratio of total COVID-19 to annual past pneumonia deaths | No. of days when daily COVID-19 deaths exceed past daily pneumonia deaths |  |  |
| --- | --- | --- | --- | --- | --- | --- | --- | --- | --- | --- | --- |
|  | Median | Lower | Upper | Median | Lower | Upper |  |  | Median | Lower | Upper |
| USA | 18-Apr | 11-Apr | 30-Apr | 4,787 | 2,428 | 13,505 | 54 | 1.6 | 66 | 50 | 85 |
| UK | 21-Apr | 13-Apr | 30-Apr | 2,441 | 1,238 | 5,644 | 36 | 1.3 | 65 | 49 | 84 |
| Italy | 29-Mar | 26-Mar | 01-Apr | 1,422 | 1,001 | 2,196 | 16 | 1.6 | 66 | 57 | 80 |
| France | 07-Apr | 03-Apr | 12-Apr | 1,433 | 966 | 2,439 | 12 | 2.1 | 63 | 52 | 78 |
| Spain | 01-Apr | 30-Mar | 05-Apr | 1,468 | 1,000 | 2,455 | 12 | 2.1 | 61 | 51 | 76 |
| Canada | 04-May | 15-Apr | 21-May | 512 | 120 | 1,973 | 7 | 1.5 | 73 | 39 | 104 |
| Belgium | 09-Apr | 05-Apr | 16-Apr | 494 | 311 | 905 | 5 | 1.8 | 64 | 51 | 80 |
| Germany | 13-Apr | 07-Apr | 04-May | 443 | 262 | 2,145 | 21 | 0.4 | 43 | 30 | 79 |
| Iran | 28-Mar | 25-Mar | 21-Apr | 298 | 205 | 573 | 7 | 0.9 | 60 | 51 | 97 |
| Netherlands | 05-Apr | 02-Apr | 11-Apr | 275 | 182 | 458 | 4 | 1.2 | 57 | 46 | 70 |
| Hubei, China | 12-Feb | 09-Feb | 15-Feb | 202 | 139 | 318 | 6 | 0.6 | 45 | 40 | 51 |
| Switzerland | 07-Apr | 02-Apr | 16-Apr | 114 | 74 | 217 | 2 | 1.4 | 62 | 49 | 83 |
| New York | 11-Apr | 08-Apr | 16-Apr | 1,303 | 805 | 2,356 | 15 | 1.5 | 58 | 47 | 71 |
| New Jersey | 14-Apr | 09-Apr | 21-Apr | 418 | 239 | 832 | 7 | 1.0 | 55 | 42 | 71 |
| Michigan | 11-Apr | 06-Apr | 25-Apr | 235 | 137 | 551 | 10 | 0.4 | 42 | 31 | 63 |
| Louisiana | 08-Apr | 03-Apr | 26-Apr | 101 | 65 | 246 | 4 | 0.4 | 41 | 30 | 68 |
| California | 14-Apr | 05-Apr | 11-May | 96 | 53 | 485 | 25 | 0.1 | 3 | 0 | 57 |
| Washington state | 10-Apr | 01-Apr | 05-May | 49 | 31 | 125 | 5 | 0.2 | 26 | 13 | 70 |
| **Sub-Total Countries** |  |  |  |  |  |  | **182** | **1.4** | **62** | **48** | **83** |
| **Sub-Total US States** |  |  |  |  |  |  | **66** | **0.6** | **38** | **27** | **67** |

A crude approximation of which jurisdictions have had some success in “flattening the curve” is shown in Figure 3, which contrasts the trajectories expressed as deaths per 1 million population in Italy, France, and Germany. All three countries reported their first case almost simultaneously, but Germany took the most aggressive steps to expand testing. Compared to the other two countries, the German peak is lower with fewer peak day deaths. Similarly, the rapid and early increase in deaths in New York state (mainly in New York City), which accounts for about 25% of the forecasted USA death totals, contrasts with the flatter trajectory in Washington and California, which both adopted reasonably aggressive efforts in physical distancing and other control measures. The upper credible interval of rates per million in Washington and California is below the lowest credible interval in New York. The contrast in cumulative rates per 1 M population across the countries with rising totals, and the US states (Figure 4) also illustrate marked divergence in mortality trajectories. New York has the highest cumulative median mortality rate per million (1135) which is about 12-fold that of Germany (96).

**Figure 3:**
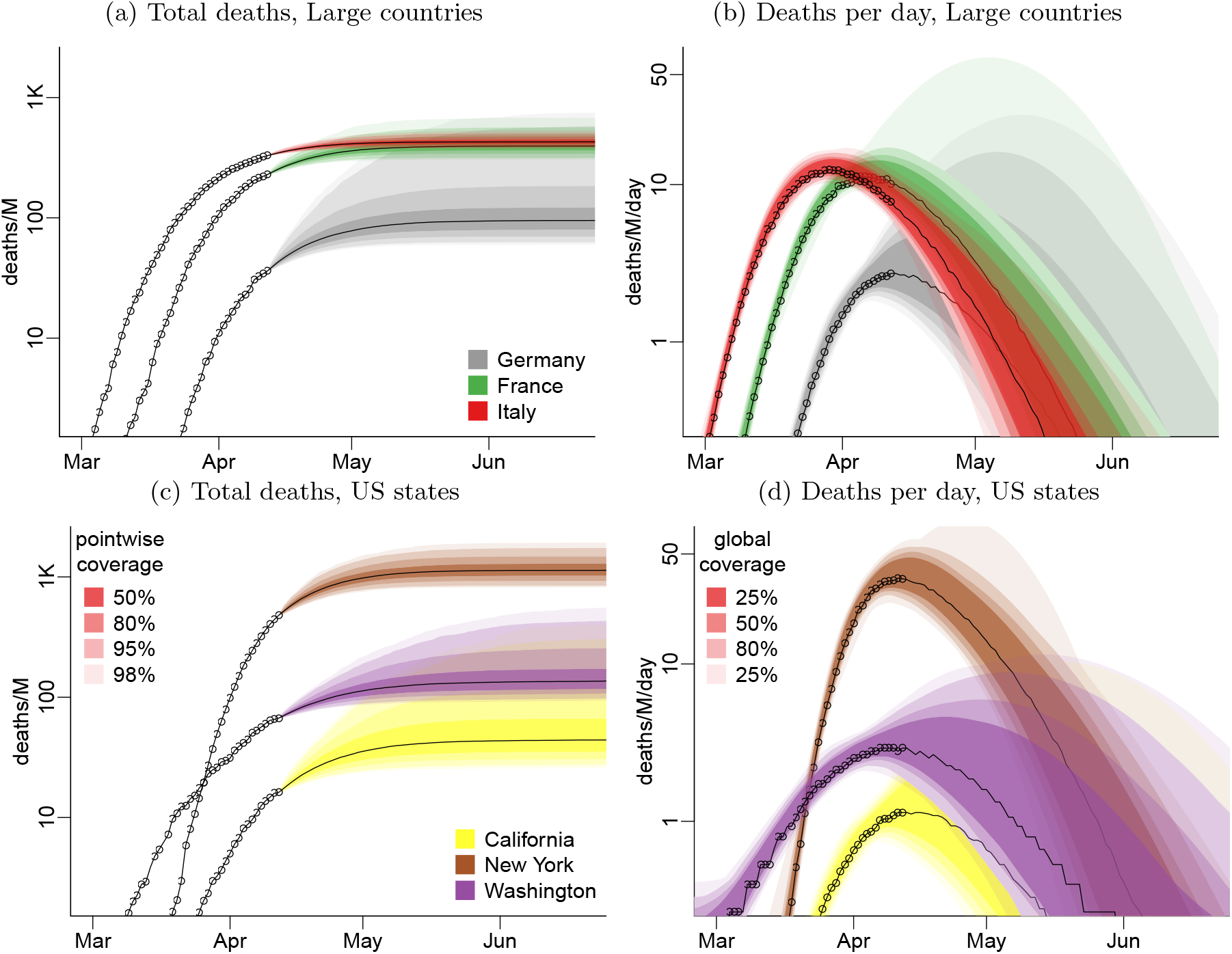
Deaths by country

**Figure 4:**
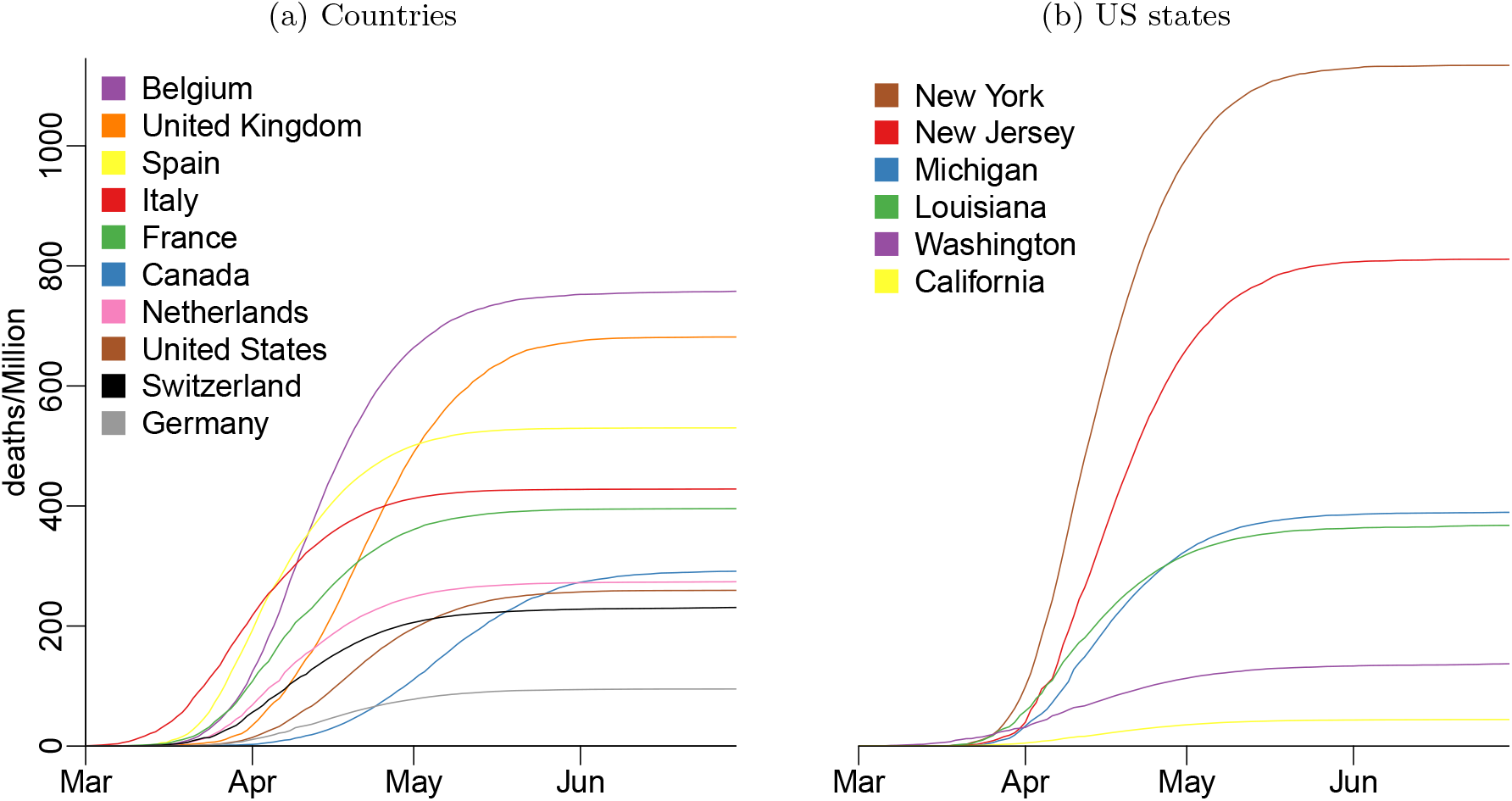
Comparison of cumulative mortality per million.

## DISCUSSION

Our forecasting predicts that in 12 early high-burden countries, the minimum number of COVID-19 deaths will be about 50% higher than current, reaching about 167,000 deaths. However, a worst case scenario would be just under 593,000 deaths, or a 500% increase over current totals. Much of the trajectory across these countries depends on what happens in the USA, which accounts for 30-40% of the cross country forecasted totals. Within the USA, COVID-19 deaths will be a minimum of 49,000 or about 2 fold over current, but could reach 249,000. Within the US, most of the increase will be from outside the initial six states we examined, and in these settings micro epidemics with different trajectories are likely to occur, as has been observed in sub-parts of Italy and Spain.^1-3^

The wide variation in forecasted mortality suggests interventions might help reduce the eventual total. Reassuringly, our analyses is sufficiently robust to fit actual death patterns in Italy and Spain, and confirms that Germany, Washington and California, which have taken testing, isolation and other strategies seriously have flatter trajectories and lower daily totals than areas with looser measures. This has allowed valuable time to scale up acute bed and ventilator capacity.

An acute excess of hospitalizations and deaths, clustered over short time periods, can swamp any health system, as indeed happened in northern Italy. Italy faced more pressures in one month on ventilator and critical-care bed supply than the usual annual burden.^21^ Model-based estimates for the USA suggest that above age 55 years, ten people will need hospitalization for every COVID-19 death.^22^ For each country, GCAM provides additional details of when these deaths will peak and the duration of excess deaths, which will average about 62 days but with a wide range (Indeed suggesting that California already has sufficient capacity to meet the surge of cases). Such information should aid planning and help avert an overwhelming of health systems.

Initial infectious disease models estimated total COVID-19 deaths in the USA of 1 to 2.2 million (roughly 40% to 80% over and above the usual annual total of 2.8 million deaths) if no action were taken.^23^ These same projections suggested 510,000 COVID-19 deaths in the UK, roughly doubling the current annual total. The Institute for Health Metrics and Evaluation (IHME) model considers physical distancing through May 2020 and has median death estimates lower than ours (Supplementary Appendix). However, the IHME model has some implausible results like suggesting that Netherlands will have 16,000 deaths which is only slightly below their estimate for France and Spain, despite being at least three times larger in population.^22^ Moreover, the IHME analyses assumes a symmetrical bell-shaped mortality curve, whereas a skewed curve (which we use) is observed in the actual data.^1-3^ Our totals for the USA fall broadly within the range projected by the White House Task Force of 100,000 to 240,000 deaths,^24^ and our Canadian estimates are consistent with infectious disease dynamic model results.^25^

There is little doubt that some of GCAM forecasts are wrong as undercounts of COVID-19 and delayed reporting are common.^4-6^ For example, the undercount in UK COVID-19 deaths might have been about 70% for some weeks.^4^ For France, Italy, the Netherlands and the UK, we could find some record of undercounts and correcting for these substantially raised the total deaths in each country (Supplementary Appendix). However, systematic examination of undercounts across settings is needed to establish if the undercounts have accelerated during the epidemic, which would mean that we have underestimated the speed of the upward trajectory, or if they are more random, in which case our results would mostly be unaffected. Future iterations of GCAM will incorporate undercounts in as many countries as document these. Differences in reporting across countries does not bias the results for any one country however, as mortality forecasts rely on actual counts within each country. Third, in settings with few deaths over many days, the model becomes highly sensitive to the assumed prior distributions. However, the model validation (Supplementary Appendix) showed reasonably good performance with actual death counts in several countries. As with all models, ours emphasizes the need to obtain additional days of observation and rerun the analyses as data improve, and as additional epidemiological research results become available.^26^ The GCAM is set up to be do so, hence its results improve constantly.

In a time of crisis, open data sharing is particularly important, hence the full input details and code of GCAM are freely available (www.cghr.org/covid) and we welcome criticisms and suggestions on how to refine our estimates. We provide a simple metric of precision that can be used to gauge improvements over time. Future iterations will also provide estimates for sub-national regions of the USA and other countries, particularly for Asian and African countries with weaker health systems and mortality surveillance capacity.^27^

## Data Availability

All data will be available on www.cghr.org/covid including the source code for the analyses.

## Article Information

### Author Contributions

Dr P Brown had full access to all of the data in the study and takes responsibility for the integrity of the data and the accuracy of the data analysis.

*Concept and design:* P Brown, Jha.

*Acquisition, analysis, or interpretation of data:* All authors.

*Drafting of the manuscript:* Jha, P Brown.

*Critical revision of the manuscript for important intellectual content:* All authors.

*Statistical analysis:* P Brown, Rai, Tang, Suraweera, Jha.

*Obtained funding:* Jha.

*Administrative, technical, or material support:* Rodriguez, Schultz, Suraweera, Tang, Newcombe.

*Supervision:* P Brown.

### Additional Contributions

We thank Vedika Jha for data entry assistance, and Richard Wen for assistance with the website for data.

